# Child diarrhea in Cambodia: A descriptive analysis of temporal and geospatial trends and logistic regression-based examination of factors associated with diarrhea in children under five years

**DOI:** 10.1101/2024.05.08.24307034

**Authors:** Samnang Um, Channnarong Phan, Sok Sakha, Leng Dany

## Abstract

Diarrhea is a global public health problem that is the third leading cause of death in under five years, with an estimated 1.7 billion cases in 2023 and 1.8 million deaths from diarrhea diseases every year. To better understand child diarrhea in Cambodia, we examined descriptively temporal and geospatial trends of diarrhea and used multivariate logistic regression to analyze its association with individual and household characteristics and diarrhea among children aged 0-59 months using data from the Cambodia Demographic and Health Survey for 2005 to 2022. The prevalence of diarrhea among children aged 0–59 months decreased from 19.7% in 2005 to 6.2% in 2022. The highest prevalence of childhood diarrhea in Kampong Cham (30.1%), in Mondul Kiri/Ratanak Kiri (24.6%), Battambang/Pailin (20.9%), and Mondul Kiri/Ratanak Kiri (12.9%) for 2005, 2010, 2014 and 2022. After adjusting for other variables, factors independently associated with childhood diarrhea included mothers aged 25-34 years (adjusted odds ratio (AOR) = 0.68; 95% CI: 0.48–0.96), 35-49 years (AOR = 0.60; 95% CI: 0.42–0.87), completed higher education (AOR = 0.61; 95% CI: 0.41–0.91), and employed (AOR = 0.79; 95% CI: 0.72– 0.96). Children were less likely to have diarrhea if they were older than 36 months, richest household (AOR = 0.69; 95% CI: 0.55–0.86), coastal region (AOR = 0.53; 95% CI: 0.41–0.69), born to smoker mothers (AOR = 1.61; 95% CI: 1.25–2.08), had barrier access to healthcare services (AOR = 1.20; 95% CI: 1.07–1.35), or children aged 6–23 months. Diarrhea remains highly prevalent among children in Cambodia. Public health interventions and policies to alleviate diarrhea should be prioritized to address these factors across geographical.

## Introduction

Diarrhea is defined by the World Health Organization (WHO) as passing loose or watery stool three or more times in 24 hours [1]. Although categorized in several ways, acute and persistent diarrhea are the most common classifications [1]. Persistent diarrhea is defined as diarrhea that starts acutely and lasts for more than 14 days [1]. An infection causes acute diarrhea and typically starts 12 hours to 4 days after exposure and goes away in 3 to 7 days [1]. Diarrhea is usually a symptom of an intestinal infection of various bacterial, viral, and parasitic organisms contained in food, drinking water, or from person to person due to improper hygiene [1]. Diarrhea is a global public health problem that is the third leading cause of death among under five years, with an estimated 1.7 billion childhood diarrhea cases in 2023 [1]. An estimated 1.8 million children under the age of five die from diarrheal illnesses every year, with over 80% of these deaths suffering in developing nations [3,4], including Southeast Asia [3,4]. In Southeast Asia, diarrheal diseases continue to rank among the top five causes of death, contributing to 6% of all deaths [2]. Even though still morbidity and mortality of children due to diarrhea are high in Cambodia, the percentage of children under five who had experienced diarrhea two weeks before the survey was 19.5% in 2005; this number slightly decreased to 15% in 2010 and 13% in 2014, and it also dramatically reduced to 6% in 2022 [3–6].

Additionally, the under-five mortality rate (U5M) in Cambodia has decreased from 124 deaths per 1,000 live births in 2000 to 16 deaths per 1,000 live births in 2022 [3–6]. However, its mortality is still unacceptably high due to various causes that are preventable. The under-5 mortality rates are higher in rural areas (15 deaths per 1,000 live births and 20 deaths per 1,000 live births, respectively) than in urban areas. Under-five mortality rates were highest in Ratanak Kiri province (43 deaths per 1,000 live births), Mondul Kiri province (43 deaths per 1,000 live births), Preah Vihear, and Kompong Chhang (each 36 deaths per 1,000 live births) [3–6]. Also, U5M is still higher than in the neighboring countries such as Vietnam and Thailand [7]. Diarrheal illness is a significant public health concern in Cambodia. It is still one of the top 10 causes of disability-adjusted life years (DALYs) across all age groups and a substantial cause of death for children [2]. Many associated factors with diarrhea included the age of children, place of residence, maternal education, and household economic condition [8–10]. Children living in households using improved drinking water, improved toilet facilities, and safe waste disposal were associated with lower odds of diarrhea [8–10].

Furthermore, breastfeeding practices, eating habits, and good handwashing practices were found to be significantly associated with diarrhea in children [8–10]. The incidence of diarrhea has been reported as compared to other developing countries, including malnutrition and early childhood mortality, which are unacceptably high in Cambodia. Furthermore, to prevent and control diarrhea in the future, it is necessary to know the trends of diarrhea over time, the geographical distributions, and associated risk factors. This enhanced understanding will help to identify sub-populations at greater risk for diarrhea and prioritize geographic areas for targeted public health interventions. To our authors’ knowledge, no published peer-reviewed studies have been conducted nationally to identify determinants of diarrhea in Cambodia over time. Therefore, this study aims to explore descriptive temporal and geographical trends of diarrhea among under-five children across 2005, 2010, 2014, and 2022 CDHS surveys, identify factors associated with diarrhea among under-five children, and identify the factors that contributed positively or negatively associated with diarrhea among under-five children. Understanding these factors further supports policy development and programs with more effective strategies and interventions in Cambodia to reduce the prevalence of diarrhea among under-five children and associated health risks such as childhood mortality.

## Methods

### Data source

We used children’s data from the 2005, 2010, 2014, and 2022 CDHS. The CDHS is a nationally representative population-based household survey conducted roughly every five years. The survey typically uses two-stage stratified cluster sampling to collect the samples from all provinces divided into urban and rural areas. In the first stage, clusters, or enumeration areas (EAs) representing the entire country, are randomly selected from the sampling frame using probability proportional to cluster size (PPS). The second stage then systematically samples households listed in each cluster or EA. Then, interviews were conducted with women aged 15-49 who gave birth five years before the survey in the selected households. Data about the maternal and children demographic, household characteristics, health status, behaviors, and access to health care were collected when the enumerators identified children in the selected households. Details of the CDHS survey reported have been described elsewhere [3–6]. We limited our analysis to live births in the last five years before the surveys and children collected on diarrhea. This resulted in a total sample size of 30,314 (**29,742 weighted**) children 0–59 months, including in the final analysis (see **Fig 1**).

**Fig 1.**
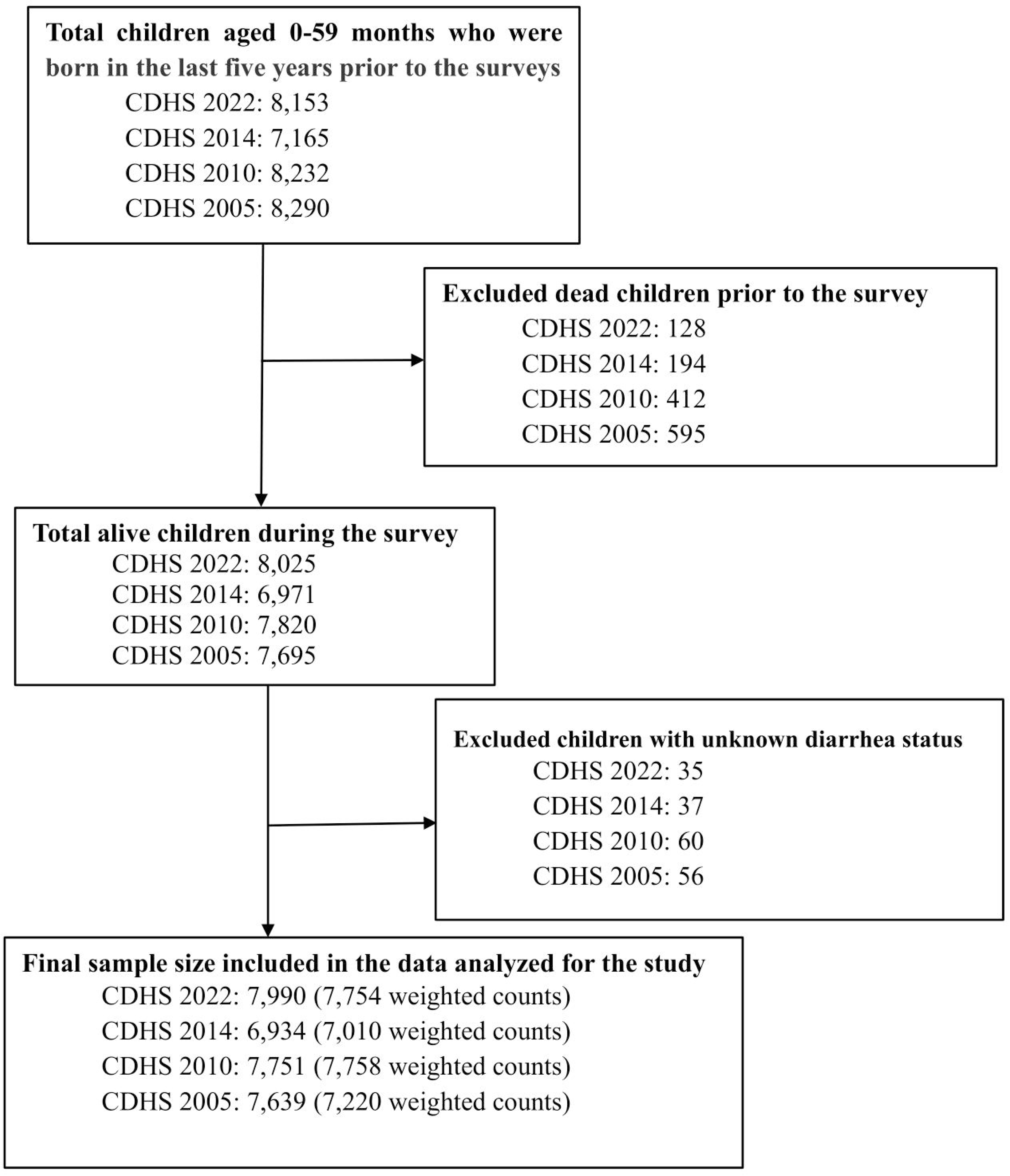
The total sample size from the 2005, 2010, 2014, and 2022 CDHS

### Measurements

#### Outcome variable

The outcome variable of this study was children under five years old having diarrhea for the last two weeks before the data collection, which was coded as “Yes = 1” and “No = 0”.

#### Indenpedent variables

**Maternal characteristics** included age (coded as 1 = 15–18 (reference), 2 = 19–24, 3 = 25–34, and 4 = 35–49 years); education (coded as 0 = no education (reference), 1 = primary, and 2 = secondary, and 3 = higher); employment (coded as 0 = not working (reference), 1 = working); smoking (coded as 0 = non-smoker and 1 = smoker); media exposure (coded as 0 = no and 1 = yes); and healthcare barriers was coded as a dichotomous variable with No Barriers = 0 and 1 or more barriers = 1 (possible barriers reported included distance, money, and waiting time).

**Child’s characteristics** included sex of child (coded as 0 = female and 1 = male); child age (coded as 0 = 0–5 (reference), 1 = 6–11, 2 = 12–23, 3 = 24–35, 4 = 36–47, and 5 = 48–60 months); birth order (coded as 1 = first child (reference), 2 = 2–3, 3 = 4–5 and 4 = 6 or more); twin birth (coded as 0 = single and 1 = twin); still breastfeeding (coded as 0 = no and 1 = yes); and received measles vaccinated (coded as 0 = no and 1 = yes); and vitamin A supplementation within the last six months (coded as 0 = no and 1 = yes).

**Household characteristics** included the DHS wealth index, coded as an ordinal level variable with richest = 1 (reference), richer = 2, middle = 3, poorer = 4, and poorest = 5 (we opted to use the wealth index that was provided by each respective survey year of the CDHS as opposed to calculating a wealth index across the pooled data. This decision was based on prior research that found wealth status designation was comparable across the CDHS-provided indices and an index based on data pooled across years) [3–6]; source of drinking water (coded as 1 = unimproved and 0 = improved), type of toilet (coded as 1 = unimproved and 0 = improved; and residence (coded as 1 = urban and 2 = rural). Cambodia’s domains/provinces were regrouped for analytic purposes into a categorical variable with five geographical regions that were coded as Phnom Penh capital city = 1 (reference), Plains = 2, Tonle Sap = 3, costal/sea = 4, and mountains = 5 (Phnom Penh capital city; Plains included, Kampong Cham/Tbong Khmum, Kandal, Prey Veng, Svay Rieng, and Takeo; Tonle Sap included Banteay Meanchey, Kampong Chhnang, Kampong Thom, Pursat, Siem Reap, Battambang, Pailin, and Otdar Meanchey; Coastal/sea included Kampot, Kep, Preah Sihanouk, and Koh Kong; and Mountains included Kampong Speu, Kratie, Preah Vihear, Stung Treng, Mondul Kiri, and Ratanak Kiri). Survey year was coded as 1 = 2005 (reference), 2 = 2010, 3 = 2014 and 4 = 2022.

### Statistical analysis

Statistical analyses were performed using STATA version 18 (Stata Crop 2023, College Station, TX) [11]. We formally incorporated the DHS’s complex sample design using the “survey” package; all estimations were carried out using the svy command in our descriptive and logistic regression analyses. Key maternal, child, and household characteristics were described using weighted frequency distributions for specific survey years and pooled together for all years. A temporal series of maps illustrating provincial variations in the prevalence of anemia over time was created using ArcGIS software version 10.3 [12]. Cambodian shapefile of provinces were obtained from the publicly accessible DHS website under a copyright license (https://spatialdata.dhsprogram.com/boundaries/#view=table&countryId=KH). Data quality assurance and minor data cleaning followed standard procedures in preparation for statistical analysis [13].

Bivariate analysis using the Chi-square test assessed associations between the variables of interest (including maternal-child, household characteristics, and geographical regions) and diarrhea status. The multivariate logistic regression analysis used variables associated with diarrhea status with a p-value ≤ 0.05 [14,15]. Women’s age, place of residence, and survey years were included in the multivariate regression based on the literature and prior knowledge [8]. Simple logistic regression was used to analyze associations between diarrhea and maternal, children, household characteristics, and geographical regions. Results are reported as unadjusted odds ratios (COR) with 95% confidence intervals (CI) and corresponding p-values. Multivariate logistic regression analysis was then used to assess independent factors associated with diarrhea after adjusting for other covariate variables in the final model. The final multivariate logistic regression results were reported as adjusted odds ratios (AOR) with 95% confidence intervals and corresponding p-values. The final multivariate logistic regression model results were considered statistically significant based on a p-value less than 0.05 and 95% confidence intervals that did not cross an AOR of 1.0. Potential multi-collinearity between right-hand-sided variables was checked using a continuous version of the outcome variable and evaluating VIF scores after fitting a regression model (**S1 Table**).

### Ethical consideration

The Cambodia Demographic and Health Survey (CDHS) 2005-2022 was approved by the Cambodia National Ethics Committee for Health Research and ICF’s Institutional Review Board (IRB) in Rockville, Maryland, USA. The study participants were informed about the survey before their interview, and for each participant under 18 years old, signed informed consent was obtained from their parents or guardians. CDHS data are publicly accessible upon request through the DHS website at (URL: https://dhsprogram.com/data/available-datasets.cfm) following permission obtained through an online request outlining the purpose of our investigation and removing all participant personal information. The details about the CDHS’s report [3–6].

## Results

The pooled data included 29,742 children aged 0–59 months; there were 7,220 in 2005, 7,758 in 2010, 7,010 in 2014, and 7,754 in 2022, respectively. Overall, 21.7% had a mother above 35 years old, 31.3% had a mother who had at least completed secondary education, 16.6% did not receive formal schooling, 37.2% had an unemployed mother, and 2.9% had a mother who smoked. 34.1% of children were first-order births, and 6.7% were sixth- or high-order births. 1.4% of children were twins, 49.5% were girls, 46.3% were still breastfeeding, 60.9% received the measles vaccine, and 49.1% were Vitamin A intake. Regarding households characterized by children, 78.8% resided in rural areas, 24.6% belonged to the poorest families, 45.8% belonged to households drinking unimproved water, and 51.0% were unimproved toilets (see **Table 1**).

**Table 1.** Descriptive statistics of individual and household socio-demographic characteristics.

| Variables | 2005 |  | 2010 |  | 2014 |  | 2022 |  | 2005-2022 |  |
| --- | --- | --- | --- | --- | --- | --- | --- | --- | --- | --- |
|  | (n=7,220) |  | (n=7,758) |  | (n=7,010) |  | (n=7,754) |  | (n=29,742) |  |
|  | Freq. | % | Freq. | % | Freq. | % | Freq. | % | Freq. | % |
| **Had diarrhea in the past two weeks** |  |  |  |  |  |  |  |  |  |  |
| Not diarrhea | 5,800 | 80.3 | 6,597 | 85.0 | 6,108 | 87.1 | 7,276 | 93.8 | 25,782 | 86.7 |
| Diarrhea | 1,420 | 19.7 | 1,161 | 15.0 | 902 | 12.9 | 477 | 6.2 | 3,960 | 13.3 |
| **Maternal Characteristic** |  |  |  |  |  |  |  |  |  |  |
| **Age (years)** |  |  |  |  |  |  |  |  |  |  |
| <19 | 89 | 1.2 | 90 | 1.2 | 92 | 1.2 | 98 | 1.3 | 370 | 1.2 |
| 19-24 | 1,918 | 26.6 | 1,825 | 23.5 | 1,828 | 23.6 | 1,457 | 18.8 | 7,029 | 23.6 |
| 25-34 | 3,321 | 46.0 | 4,282 | 55.2 | 4,022 | 51.8 | 4,269 | 55.0 | 15,894 | 53.4 |
| 35-49 | 1,891 | 26.2 | 1,561 | 20.1 | 1,068 | 13.8 | 1,930 | 24.9 | 6,450 | 21.7 |
| **Educational** |  |  |  |  |  |  |  |  |  |  |
| No education | 1,720 | 23.8 | 1,419 | 18.3 | 962 | 12.4 | 841 | 10.8 | 4,942 | 16.6 |
| Primary | 4,251 | 58.9 | 4,385 | 56.5 | 3,675 | 47.4 | 3,183 | 41.0 | 15,494 | 52.1 |
| Secondary | 1,207 | 16.7 | 1,833 | 23.6 | 2,175 | 28.0 | 3,160 | 40.7 | 8,375 | 28.2 |
| Higher | 42 | 0.6 | 120 | 1.5 | 198 | 2.6 | 570 | 7.3 | 931 | 3.1 |
| **Employment** |  |  |  |  |  |  |  |  |  |  |
| Not working | 2,952 | 40.9 | 2,629 | 33.9 | 2,523 | 36.0 | 2,958 | 38.1 | 11,061 | 37.2 |
| Working | 4,267 | 59.1 | 5,130 | 66.1 | 4,483 | 64.0 | 4,796 | 61.9 | 18,676 | 62.8 |
| **Smoking** |  |  |  |  |  |  |  |  |  |  |
| No-smoker | 6,878 | 95.3 | 7,546 | 97.3 | 6,813 | 97.2 | 7,653 | 98.7 | 28,890 | 97.1 |
| Smoker | 342 | 4.7 | 212 | 2.7 | 197 | 2.8 | 100 | 1.3 | 852 | 2.9 |
| **Media exposure** |  |  |  |  |  |  |  |  |  |  |
| No | 6,846 | 94.8 | 7,427 | 95.7 | 6,808 | 97.1 | 7,654 | 98.7 | 28,736 | 96.6 |
| Yes | 374 | 5.2 | 331 | 4.3 | 202 | 2.9 | 99 | 1.3 | 1,006 | 3.4 |
| **Healthcare Barriers** |  |  |  |  |  |  |  |  |  |  |
| No barrier | 1,084 | 15.0 | 2,181 | 28.1 | 1,753 | 25.0 | 3,160 | 40.8 | 8,178 | 27.5 |
| One or More Barriers | 6,136 | 85.0 | 5,578 | 71.9 | 5,257 | 75.0 | 4,594 | 59.2 | 21,564 | 72.5 |
| **Child Characteristics** |  |  |  |  |  |  |  |  |  |  |
| **Age(months)** |  |  |  |  |  |  |  |  |  |  |
| 0-5m | 743 | 10.3 | 710 | 9.2 | 732 | 10.4 | 760 | 9.8 | 2,946 | 9.9 |
| 6-11m | 769 | 10.7 | 825 | 10.6 | 759 | 10.8 | 809 | 10.4 | 3,163 | 10.6 |
| 12-23m | 1,513 | 21.0 | 1,608 | 20.7 | 1,457 | 20.8 | 1,642 | 21.2 | 6,220 | 20.9 |
| 24-35m | 1,411 | 19.5 | 1,601 | 20.6 | 1,360 | 19.4 | 1,496 | 19.3 | 5,866 | 19.7 |
| 36-47m | 1,409 | 19.5 | 1,517 | 19.6 | 1,332 | 19.0 | 1,519 | 19.6 | 5,777 | 19.4 |
| 48-60m | 1,374 | 19.0 | 1,498 | 19.3 | 1,370 | 19.5 | 1,528 | 19.7 | 5,770 | 19.4 |
| **Sex** |  |  |  |  |  |  |  |  |  |  |
| Boys | 3,584 | 49.6 | 4,006 | 51.6 | 3,506 | 50.0 | 3,935 | 50.7 | 15,031 | 50.5 |
| Girls | 3,636 | 50.4 | 3,752 | 48.4 | 3,504 | 50.0 | 3,819 | 49.3 | 14,711 | 49.5 |
| **Birth order** |  |  |  |  |  |  |  |  |  |  |
| 1 child | 2,008 | 27.8 | 2,664 | 34.3 | 2,734 | 39.0 | 2,747 | 35.4 | 10,152 | 34.1 |
| 2-3 child | 2,866 | 39.7 | 3,365 | 43.4 | 3,185 | 45.4 | 4,172 | 53.8 | 13,588 | 45.7 |
| 4-5 child | 1,389 | 19.2 | 1,117 | 14.4 | 801 | 11.4 | 699 | 9.0 | 4,006 | 13.5 |
| ≥ 6 child | 958 | 13.3 | 613 | 7.9 | 290 | 4.1 | 135 | 1.7 | 1,996 | 6.7 |
| **Twin** |  |  |  |  |  |  |  |  |  |  |
| Single | 7,129 | 98.7 | 7,649 | 98.6 | 6,889 | 98.3 | 7,672 | 98.9 | 29,339 | 98.6 |
| Multiple | 91 | 1.3 | 109 | 1.4 | 121 | 1.7 | 82 | 1.1 | 403 | 1.4 |
| **Still breastfeeding** |  |  |  |  |  |  |  |  |  |  |
| No | 3,153 | 43.7 | 3,898 | 50.2 | 3,836 | 54.7 | 5,091 | 65.7 | 15,978 | 53.7 |
| Yes | 4,067 | 56.3 | 3,860 | 49.8 | 3,174 | 45.3 | 2,663 | 34.3 | 13,763 | 46.3 |
| **Measles' vaccine** |  |  |  |  |  |  |  |  |  |  |
| Not vaccinated | 2,621 | 36.3 | 2,349 | 30.3 | 1,999 | 28.5 | 4,660 | 60.1 | 11,629 | 39.1 |
| Vaccinated | 4,599 | 63.7 | 5,409 | 69.7 | 5,011 | 71.5 | 3,094 | 39.9 | 18,113 | 60.9 |
| **Vitamin A supplement** |  |  |  |  |  |  |  |  |  |  |
| No | 4,836 | 67.0 | 2,771 | 35.7 | 2,560 | 36.5 | 4,960 | 64.0 | 15,127 | 50.9 |
| Yes | 2,383 | 33.0 | 4,987 | 64.3 | 4,451 | 63.5 | 2,794 | 36.0 | 14,615 | 49.1 |
| **Household Characteristics** |  |  |  |  |  |  |  |  |  |  |
| **Wealth index** |  |  |  |  |  |  |  |  |  |  |
| Poorest | 1,930 | 26.7 | 2,035 | 26.2 | 1,684 | 24.0 | 1,668 | 21.5 | 7,317 | 24.6 |
| Poorer | 1,639 | 22.7 | 1,657 | 21.4 | 1,401 | 20.0 | 1,445 | 18.6 | 6,141 | 20.6 |
| Middle | 1,273 | 17.6 | 1,417 | 18.3 | 1,331 | 19.0 | 1,439 | 18.6 | 5,460 | 18.4 |
| Richer | 1,171 | 16.2 | 1,355 | 17.5 | 1,210 | 17.3 | 1,654 | 21.3 | 5,390 | 18.1 |
| Richest | 1,206 | 16.7 | 1,295 | 16.7 | 1,384 | 19.7 | 1,549 | 20.0 | 5,434 | 18.3 |
| **Place of residence** |  |  |  |  |  |  |  |  |  |  |
| Urban | 1,017 | 14.1 | 1,232 | 15.9 | 1,014 | 14.5 | 3,048 | 39.3 | 6,310 | 21.2 |
| Rural | 6,203 | 85.9 | 6,527 | 84.1 | 5,996 | 85.5 | 4,706 | 60.7 | 23,432 | 78.8 |
| **Region** |  |  |  |  |  |  |  |  |  |  |
| Phnom Penh | 580 | 8.0 | 622 | 8.0 | 603 | 8.6 | 1,131 | 14.6 | 2,936 | 9.9 |
| Plain | 2,766 | 38.3 | 3,068 | 39.5 | 2,580 | 36.8 | 2,576 | 33.2 | 10,990 | 37.0 |
| Tonle Sap | 2,381 | 33.0 | 2,509 | 32.3 | 2,251 | 32.1 | 2,504 | 32.3 | 9,644 | 32.4 |
| Coastal | 548 | 7.6 | 503 | 6.5 | 433 | 6.2 | 458 | 5.9 | 1,942 | 6.5 |
| Mountain | 945 | 13.1 | 1,056 | 13.6 | 1,144 | 16.3 | 1,085 | 14.0 | 4,229 | 14.2 |
| **Drinking water** |  |  |  |  |  |  |  |  |  |  |
| Improved | 3,755 | 52.0 | 4,222 | 54.4 | 3,516 | 50.2 | 4,629 | 59.7 | 16,122 | 54.2 |
| Unimproved | 3,464 | 48.0 | 3,536 | 45.6 | 3,494 | 49.8 | 3,125 | 40.3 | 13,619 | 45.8 |
| **Toilet facility** |  |  |  |  |  |  |  |  |  |  |
| Improved | 1,567 | 21.7 | 2,772 | 35.7 | 3,488 | 49.8 | 6,748 | 87.0 | 14,574 | 49.0 |
| Non-Improved | 5,653 | 78.3 | 4,986 | 64.3 | 3,522 | 50.2 | 1,006 | 13.0 | 15,167 | 51.0 |

Notes: Survey weights applied to obtain weighted percentages across survey years and pooled data. \*Phnom Penh capital city; Plains: Kampong Cham, Tbong Khmum, Kandal, Prey Veng, Svay Rieng, and Takeo; Tonle Sap: Banteay Meanchey, Kampong Chhnang, Kampong Thom, Pursat, Siem Reap, Battambang, Pailin, and Otdar Meanchey; Coastal/sea: Kampot, Kep, Preah Sihanouk, and Koh Kong; Mountains: Kampong Speu, Kratie, Preah Vihear, Stung Treng, Mondul Kiri, and Ratanak Kiri.

The overall prevalence trend of children aged 0–59 months having diarrhea in the previous two weeks significantly declined from 19.7% in 2005 to 15.0% in 2010 to 12.9%% in 2014 and decreased to 6.2% in 2022 (see **Table 1**). When stratified by age, the diarrhea trend peaked at around 30% among children aged 6–23 months in 2005. However, this trend among this age group gradually declined to less than 24% in 2010, 17% in 2014, and 12% in 2022 (see **Fig 2**).

**Fig 2.**
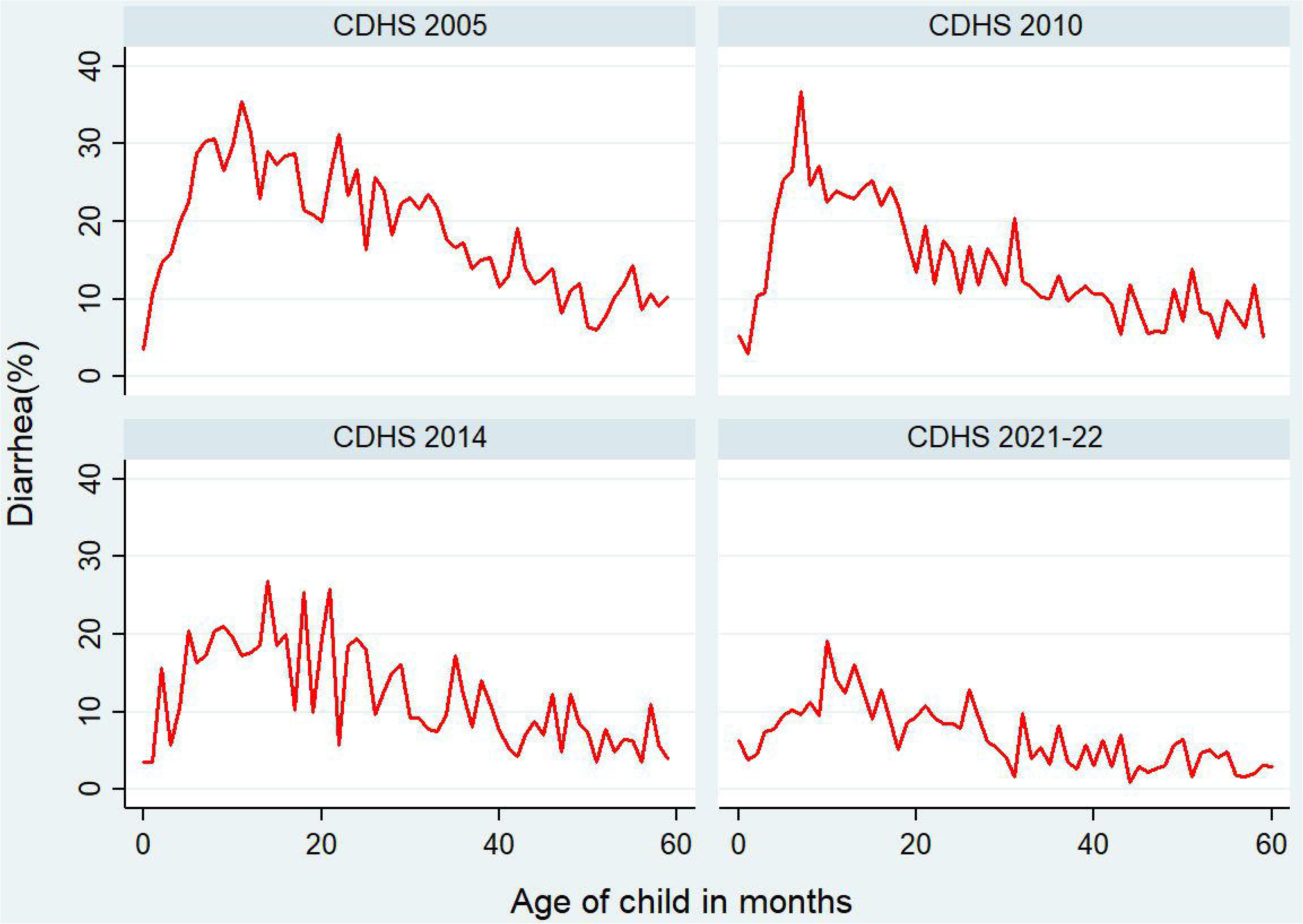
Age distribution trends of diarrhea among children 0–59 months in the last two weeks by survey years, CDHS 2000 to 2022. Notes: Survey weights were applied to obtain weighted percentages across survey years and pooled data.

The extent of this downward trend varied across the five geographical regions; the highest prevalence of diarrhea was in the Plain region in 2005 (21.7%). The Phnom Penh capital had the lowest prevalence of diarrhea across all survey years, with prevalence levels in 2022 that were lower than those of other regions (see **Fig 3**). Diarrhea was highest among children in Kampong Cham (30.1%), in Mondul Kiri/Ratanak Kiri (24.6%), in Battambang/Pailin (20.9%), and Mondul Kiri/Ratanak Kiri (12.9%) for 2005, 2010, 2014 and 2022, respectively (see **Fig 4**). The lowest prevalence of diarrhea among children was observed in Preah Sihanouk and Koh Kong provinces (9.1%), in Kampong Speu (7.5%), in Prey Veng (4.7%), and in Otdar Meanchey (1%) for 2005, 2010, 2014 and 2022.

**Fig 3.**
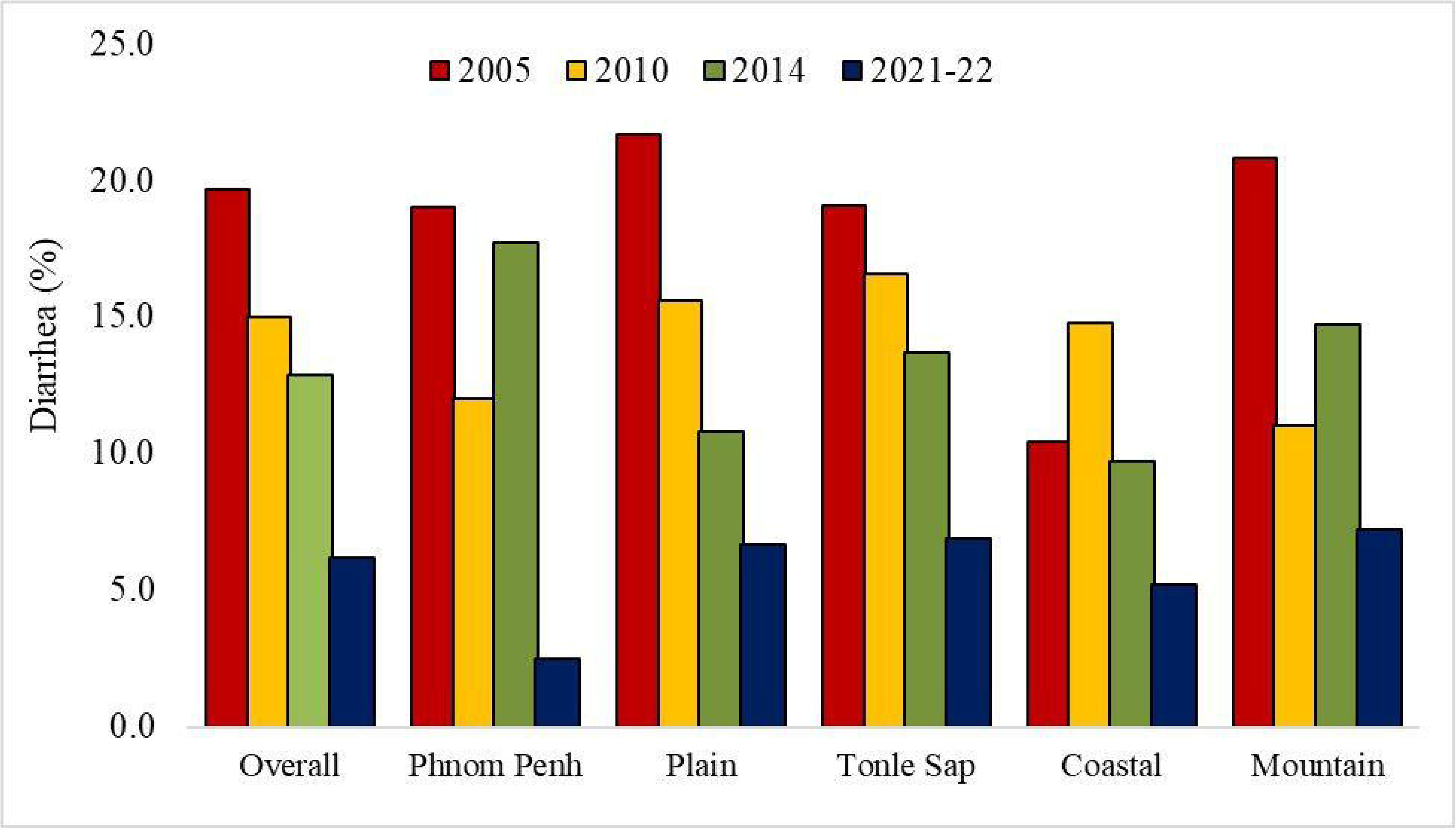
Overall and regional trends of diarrhea among children 0–59 months in the last two weeks by survey years, CDHS 2000 to 2022. Notes: Survey weights applied to obtain weighted percentages across survey years and pooled data.

**Fig 4.**
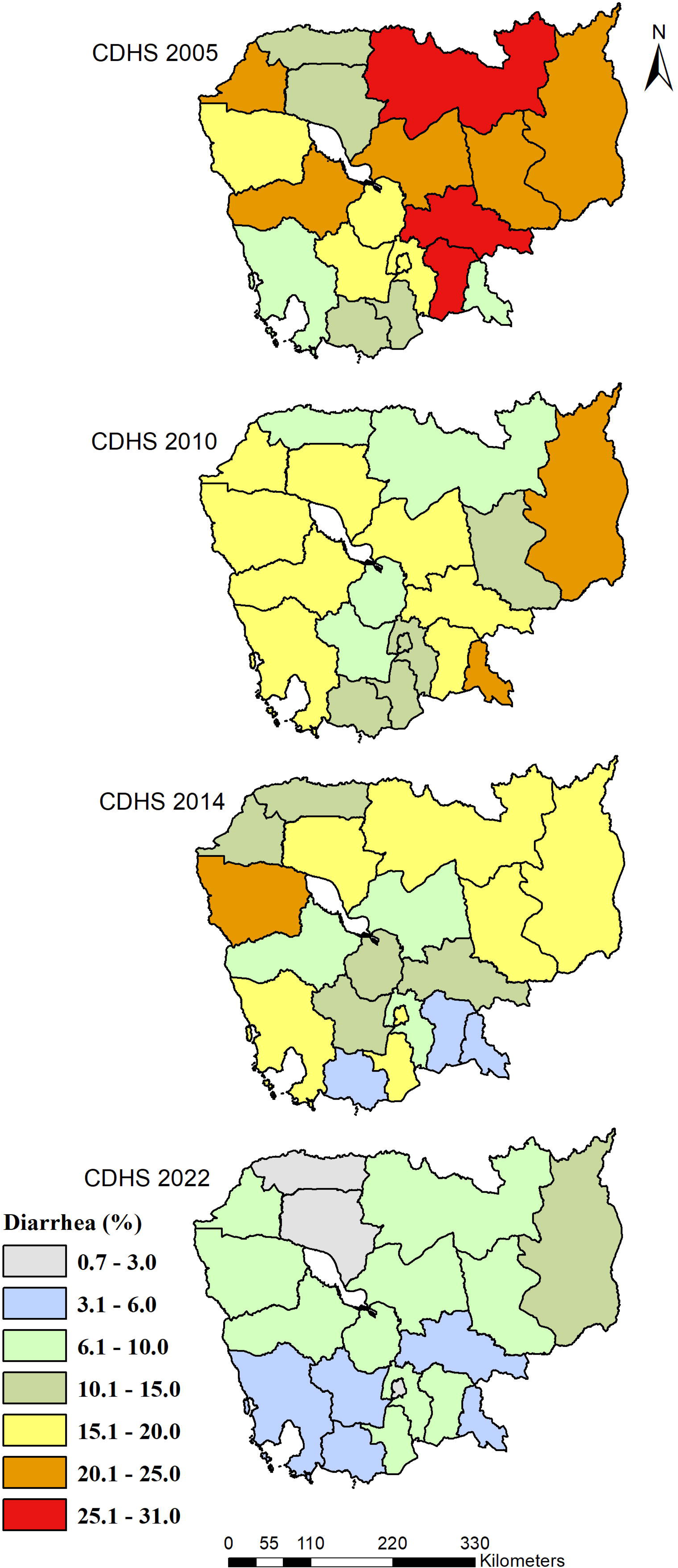
Geographical distribution of diarrhea among children 0–59 months in the last two weeks by survey years, CDHS 2000 to 2022. Cambodian shapefile of provinces were obtained from the publicly accessible DHS website under a copyright license (https://spatialdata.dhsprogram.com/boundaries/#view=table&countryId=KH). Notes: Survey weights applied to obtain weighted percentages across survey years and pooled data.

Preliminary analyses included the evaluation of cross-tabular frequency distributions combined with Chi-square tests (see **Table 2**). Maternal age was significantly associated with the risk of diarrhea in children in all survey years; the most significant percentage of children with diarrhea were those with younger mothers aged less than 19 years (25.8% in 2005, 27.8% in 2010, 20.7% in 2014 and 13.3%). Maternal educational attainment was significantly associated with diarrhea in children in all survey years; the most significant percentage of children with diarrhea were generally those whose mothers had no formal education (21.5% in 2005, 17.5% in 2010, 13.4% in 2014, and 6.3% in 2022). Maternal unemployment was also significantly associated with diarrhea in children in survey years 2005, 2014, and 2022; the prevalence of diarrhea among children who had unemployed mothers was 21.5% in 2005, 15.2% in 2014, and 7.6% in 2022. Maternal smoking of cigarettes was also significantly associated with diarrhea in children in survey years 2010 and 2014; the prevalence of diarrhea among children who had mothers smoking cigarettes was 25.0% in 2010 and 23.9% in 2014. Maternal who had at least one barrier access to healthcare were also significantly associated with diarrhea in children in all surveys; the most significant percentage of children with diarrhea were generally those whose mothers had at least one barrier access to healthcare (20.5% in 2005, 15.8% in 2010, 13.4% in 2014, and 7.0% in 2022). Diarrhea was related to children’s ages, with a higher prevalence in the age group of 6 to 11 months (6–11 months: 32% in 2005, 26.5% in 2010, 20% in 2014 and 11.6% in 2022; 12–23 months: 28.0%, 21.1%, 19.0% and 9.4%, respectively). In addition, diarrhea among boys was higher than among girls: 22.0% in 2005 and 16.1% in 2014. Breastfeeding children had a higher prevalence of diarrhea (21.8% in 2005, 17.7% in 2010, 14.2% in 2014, and 8.7% in 2022). Measles vaccination was also associated with lower diarrhea in children at 18.5% in 2005, 18.6% in 2010, and 14.2% in 2014. The proportion of children with diarrhea in rural areas was higher than in urban areas: 20.2% in 2005, 15.8% in 2010, and 7.2% in 2022. The proportion of children with diarrhea was lowest among children in the Coastal region in the survey years 2005, 2010, and 2014: 10.4% in 2005, 14.9% in 2010, 9.7% in 2014, and Phnom Penh capital city was 2.5% in 2022, respectively. Diarrhea was also associated with household toilet facilities in survey years 2005 and 2010; 21.1% in 2005 and 16.6% in 2010 of children in households that reported having non-improved toilet facilities were diarrhea. The proportion of children with diarrhea was highest among children in the poorest households across all the survey years: 22.6% in 2005, 18.5% in 2010, 16.1% in 2014, and 8.2% in 2022, respectively.

**Table 2.**
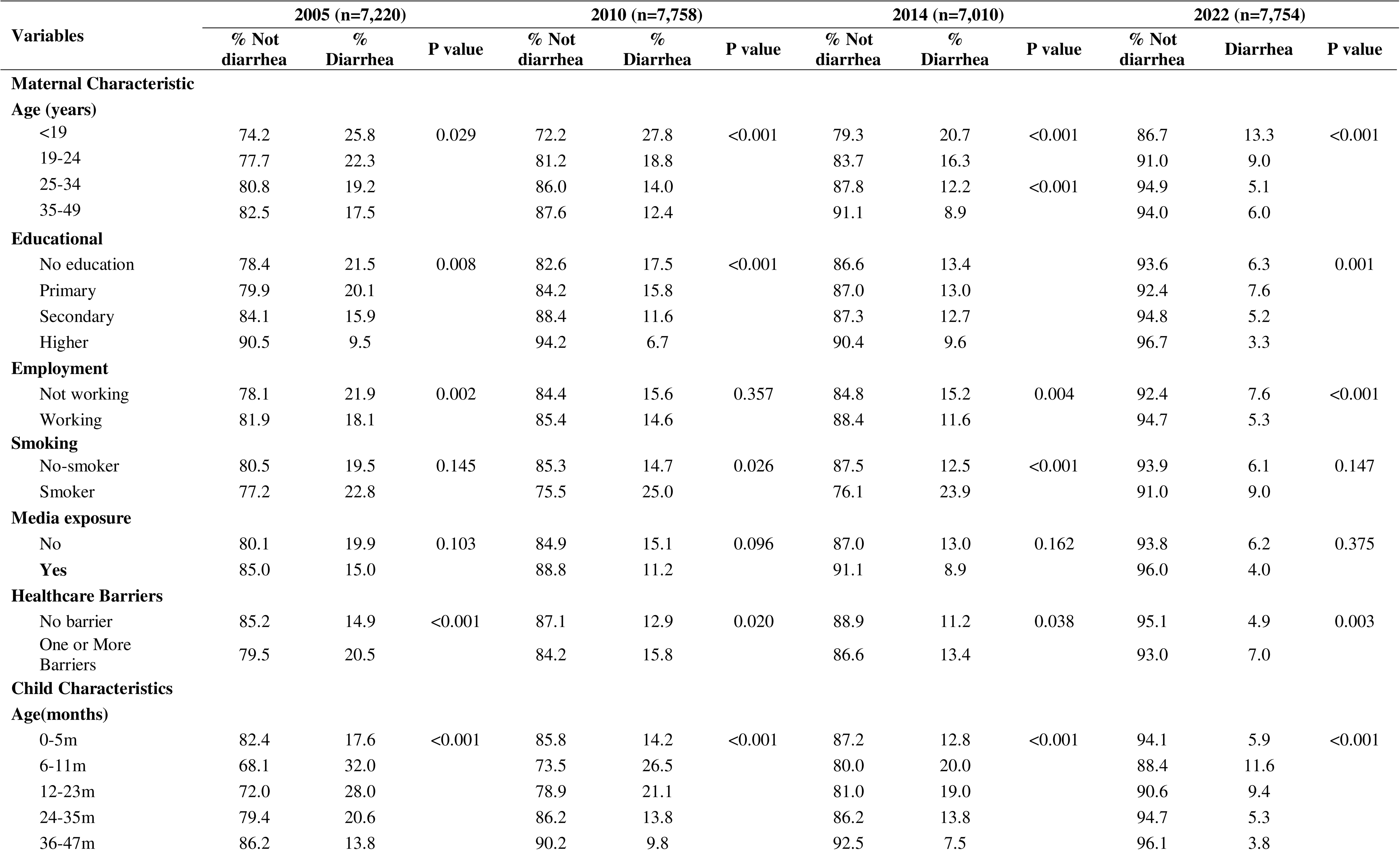
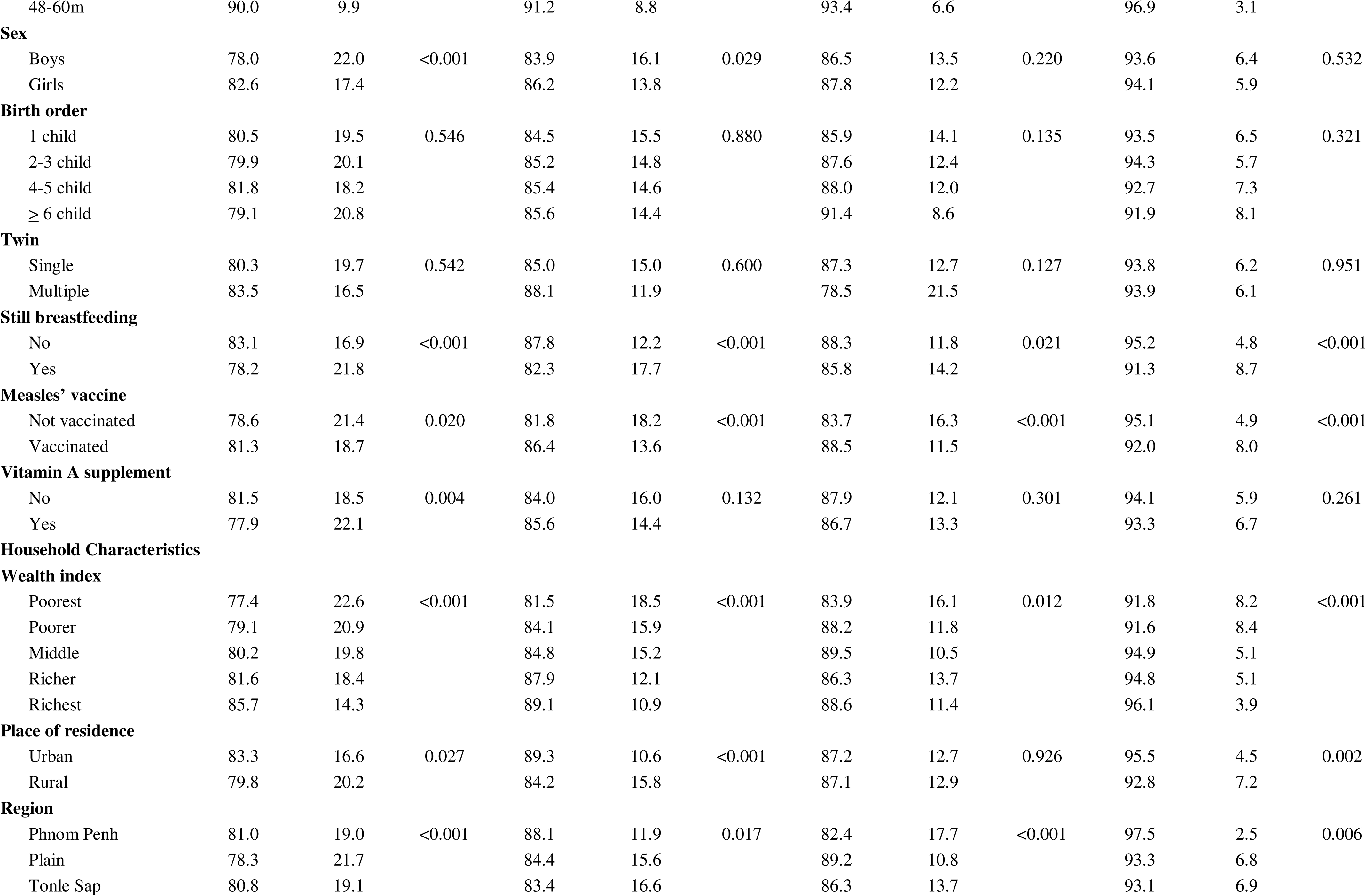
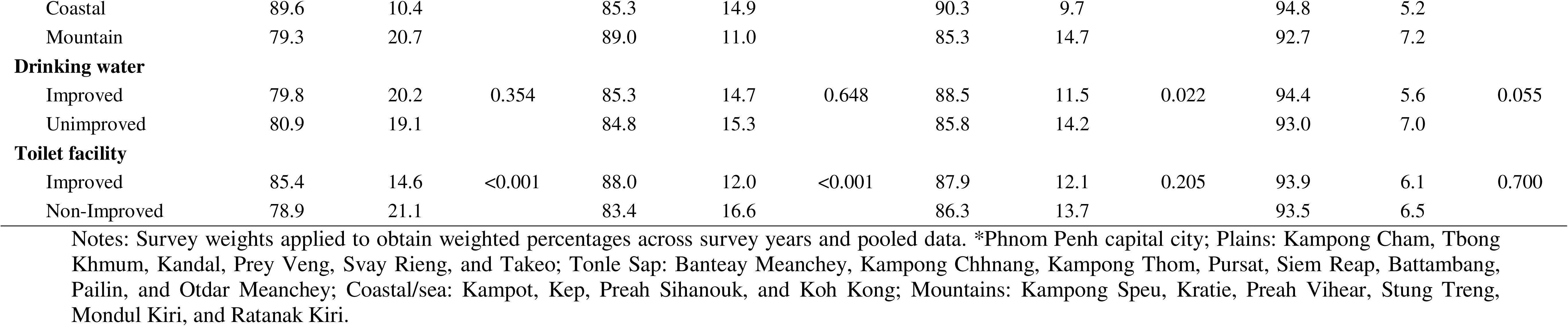
Chi-square test results for associations between child diarrhea and child, maternal, and household characteristics.

As shown in **Table 3**, several factors were independently associated with children’s decreased odds of having diarrhea in the last two weeks. These factors included children born to mothers aged 25-34 years (AOR = 0.68; 95% CI: 0.48–0.96), 35-49 years (AOR = 0.60; 95% CI: 0.42–0.87), born to mothers with secondary education (AOR = 0.84; 95% CI: 0.72–0.98), higher educated (AOR = 0.61; 95% CI: 0.41–0.91), employed mothers (AOR = 0.79; 95% CI: 0.72–0.96). For age, children aged 36–47 months (AOR = 0.64; 95% CI: 0.52–0.79) and 48–60 months (AOR = 0.52; 95% CI: 0.42–0.66) were less likely to be diarrhea than those aged 0–5 months. Also, children had lower odds of having diarrhea if they were from middle households (AOR = 0.77; 95% CI: 0.67–0.89), richer households (AOR = 0.79; 95% CI: 0.66–0.93), richest households (AOR = 0.69; 95% CI: 0.55–0.86). In addition, children had lower odds of having diarrhea if they were from the coastal region (AOR = 0.53; 95% CI: 0.41–0.69), mountain region (AOR = 0.67; 95% CI: 0.53–0.85), survey years in 2010 (AOR = 0.74; 95% CI: 0.64–0.85), 2014 (AOR = 0.62; 95% CI: 0.54–0.72), 2022 (AOR = 0.32; 95% CI: 0.26–0.38). However, children were more likely to have diarrhea if they were born to smoker mothers (AOR = 1.61; 95% CI: 1.25–2.08) compared to non-smokers. Children born to mothers who reported having at least one barrier access to healthcare service (AOR = 1.20; 95% CI: 1.07–1.35) compared to those without and children aged 6–11 months (AOR = 2.05; 95% CI: 1.66–2.52), 12–23 months (AOR = 1.68; 95% CI: 1.39–2.04).

**Table 3.**
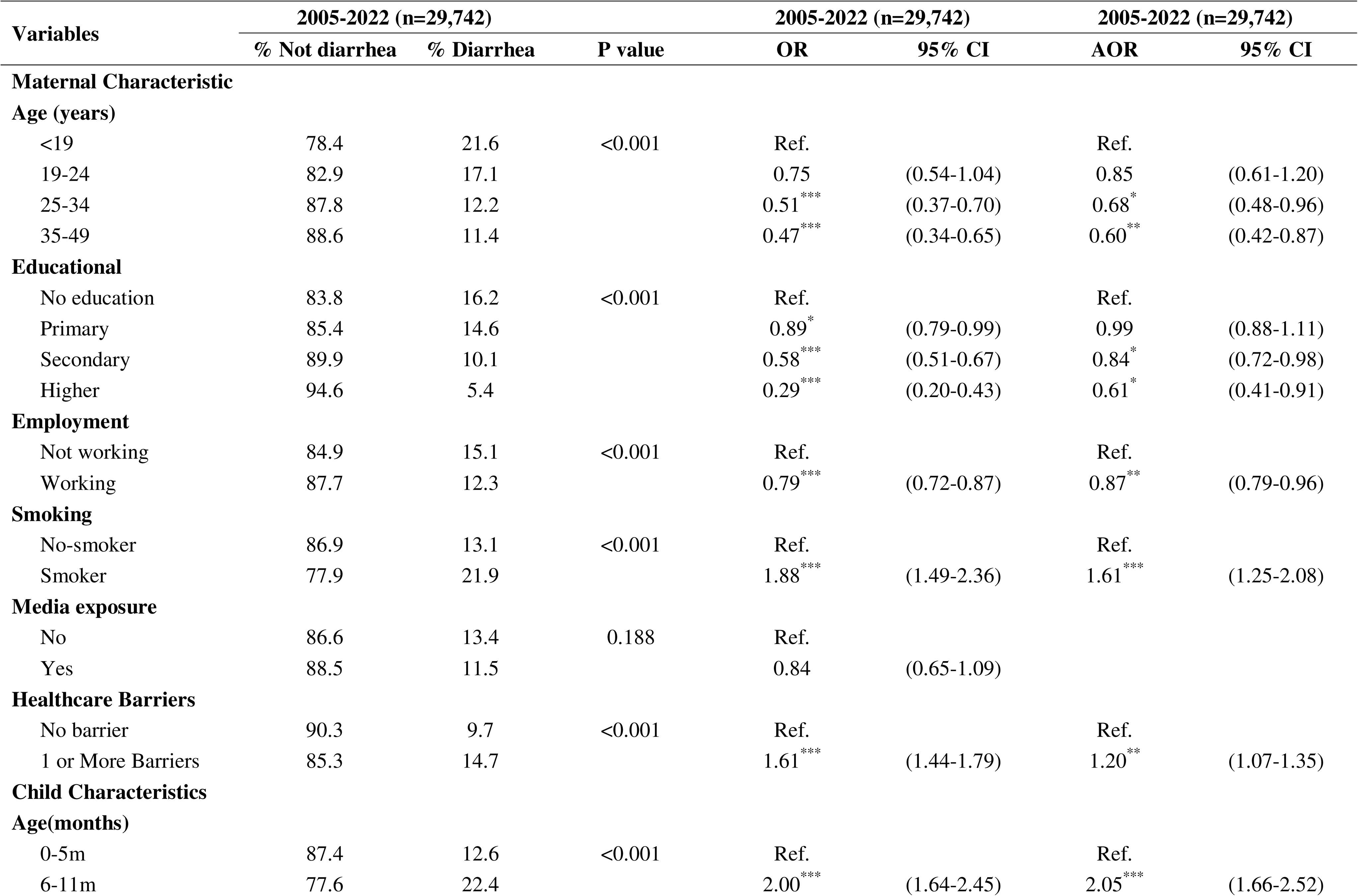
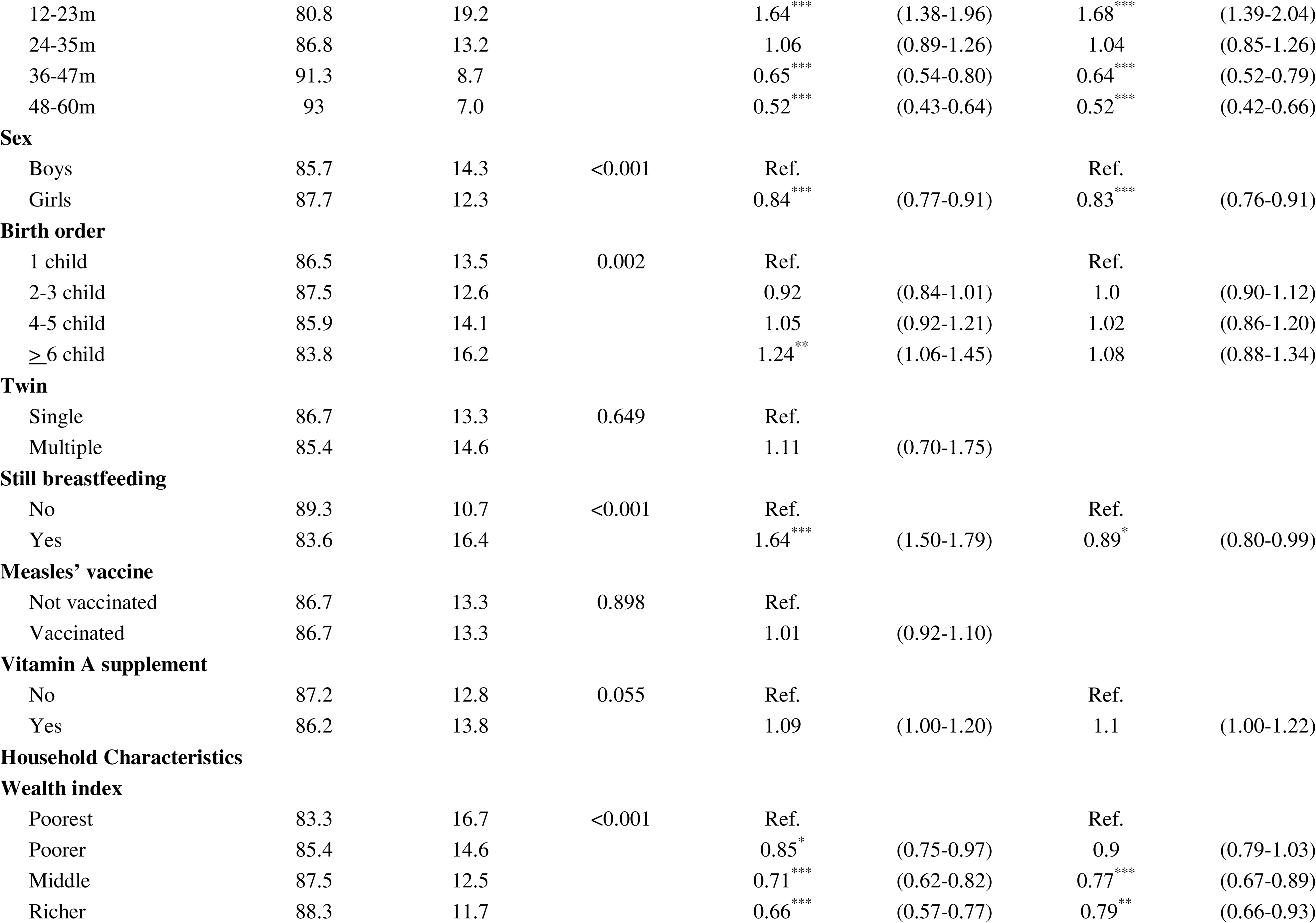
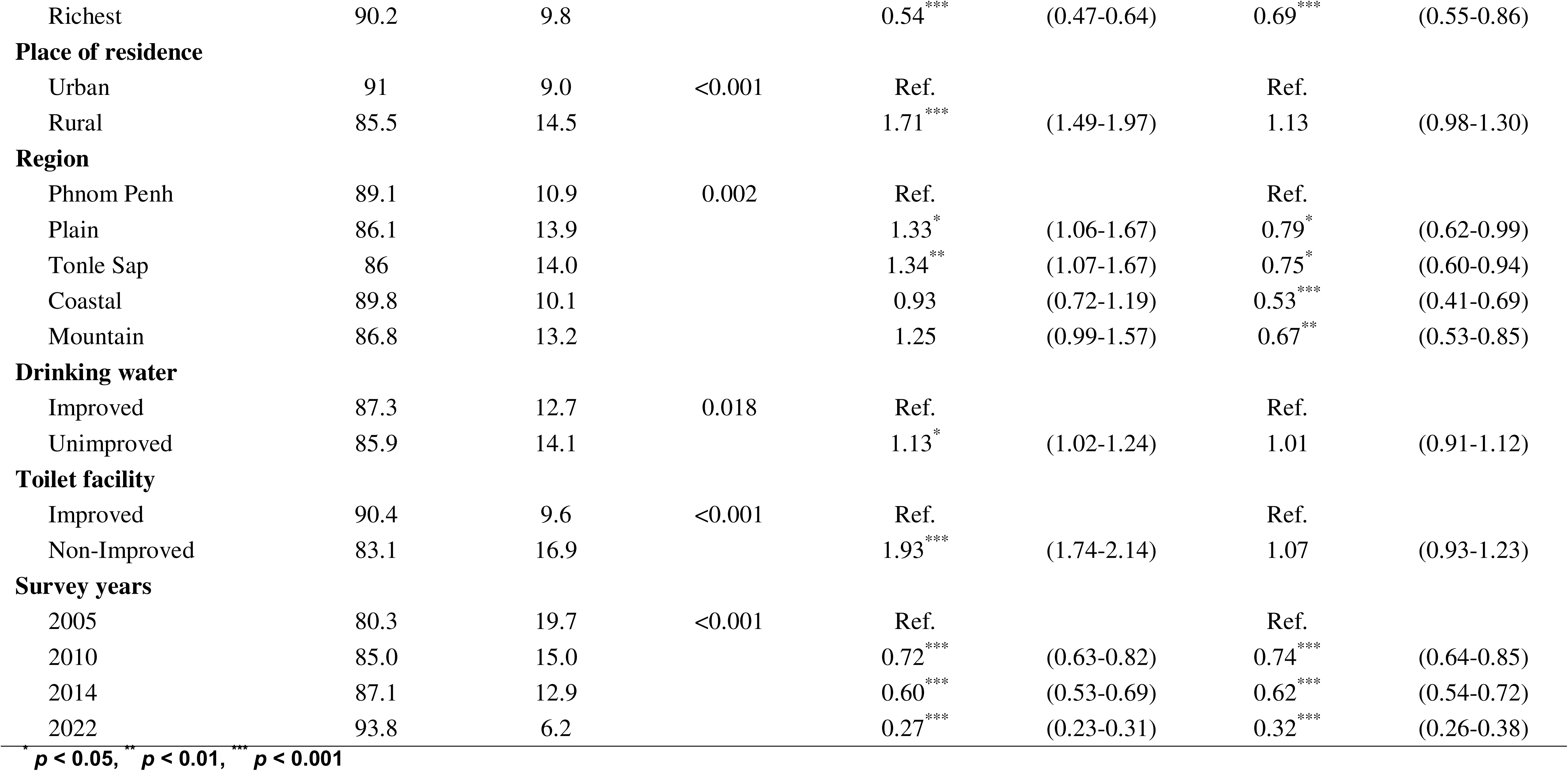
Unadjusted and adjusted analyses of child diarrhea in Cambodia.

| Variables | 2005-2022 (n=29,742) |  | P value | 2005-2022 (n=29,742) |  | 2005-2022 (n=29,742) |  |
| --- | --- | --- | --- | --- | --- | --- | --- |
|  | % Not diarrhea | % Diarrhea |  | OR | 95% CI | AOR | 95% CI |
| Maternal Characteristic |  |  |  |  |  |  |  |
| Age (years) |  |  |  |  |  |  |  |
| <19 | 78.4 | 21.6 | <0.001 | Ref. |  | Ref. |  |
| 19-24 | 82.9 | 17.1 |  | 0.75 | (0.54-1.04) | 0.85 | (0.61-1.20) |
| 25-34 | 87.8 | 12.2 |  | 0.51*** | (0.37-0.70) | 0.68* | (0.48-0.96) |
| 35-49 | 88.6 | 11.4 |  | 0.47*** | (0.34-0.65) | 0.60** | (0.42-0.87) |
| Educational |  |  |  |  |  |  |  |
| No education | 83.8 | 16.2 | <0.001 | Ref. |  | Ref. |  |
| Primary | 85.4 | 14.6 |  | 0.89* | (0.79-0.99) | 0.99 | (0.88-1.11) |
| Secondary | 89.9 | 10.1 |  | 0.58*** | (0.51-0.67) | 0.84* | (0.72-0.98) |
| Higher | 94.6 | 5.4 |  | 0.29*** | (0.20-0.43) | 0.61* | (0.41-0.91) |
| Employment |  |  |  |  |  |  |  |
| Not working | 84.9 | 15.1 | <0.001 | Ref. |  | Ref. |  |
| Working | 87.7 | 12.3 |  | 0.79*** | (0.72-0.87) | 0.87** | (0.79-0.96) |
| Smoking |  |  |  |  |  |  |  |
| No-smoker | 86.9 | 13.1 | <0.001 | Ref. |  | Ref. |  |
| Smoker | 77.9 | 21.9 |  | 1.88*** | (1.49-2.36) | 1.61*** | (1.25-2.08) |
| Media exposure |  |  |  |  |  |  |  |
| No | 86.6 | 13.4 | 0.188 | Ref. |  |  |  |
| Yes | 88.5 | 11.5 |  | 0.84 | (0.65-1.09) |  |  |
| Healthcare Barriers |  |  |  |  |  |  |  |
| No barrier | 90.3 | 9.7 | <0.001 | Ref. |  | Ref. |  |
| 1 or More Barriers | 85.3 | 14.7 |  | 1.61*** | (1.44-1.79) | 1.20** | (1.07-1.35) |
| Child Characteristics |  |  |  |  |  |  |  |
| Age(months) |  |  |  |  |  |  |  |
| 0-5m | 87.4 | 12.6 | <0.001 | Ref. |  | Ref. |  |
| 6-11m | 77.6 | 22.4 |  | 2.00*** | (1.64-2.45) | 2.05*** | (1.66-2.52) |
| 12-23m | 80.8 | 19.2 |  | 1.64<sup>***</sup> | (1.38-1.96) | 1.68<sup>***</sup> | (1.39-2.04) |
| 24-35m | 86.8 | 13.2 |  | 1.06 | (0.89-1.26) | 1.04 | (0.85-1.26) |
| 36-47m | 91.3 | 8.7 |  | 0.65<sup>***</sup> | (0.54-0.80) | 0.64<sup>***</sup> | (0.52-0.79) |
| 48-60m | 93 | 7.0 |  | 0.52<sup>***</sup> | (0.43-0.64) | 0.52<sup>***</sup> | (0.42-0.66) |
| **Sex** |  |  |  |  |  |  |  |
| Boys | 85.7 | 14.3 | <0.001 | Ref. |  | Ref. |  |
| Girls | 87.7 | 12.3 |  | 0.84<sup>***</sup> | (0.77-0.91) | 0.83<sup>***</sup> | (0.76-0.91) |
| **Birth order** |  |  |  |  |  |  |  |
| 1 child | 86.5 | 13.5 | 0.002 | Ref. |  | Ref. |  |
| 2-3 child | 87.5 | 12.6 |  | 0.92 | (0.84-1.01) | 1.0 | (0.90-1.12) |
| 4-5 child | 85.9 | 14.1 |  | 1.05 | (0.92-1.21) | 1.02 | (0.86-1.20) |
| ≥ 6 child | 83.8 | 16.2 |  | 1.24<sup>**</sup> | (1.06-1.45) | 1.08 | (0.88-1.34) |
| **Twin** |  |  |  |  |  |  |  |
| Single | 86.7 | 13.3 | 0.649 | Ref. |  |  |  |
| Multiple | 85.4 | 14.6 |  | 1.11 | (0.70-1.75) |  |  |
| **Still breastfeeding** |  |  |  |  |  |  |  |
| No | 89.3 | 10.7 | <0.001 | Ref. |  | Ref. |  |
| Yes | 83.6 | 16.4 |  | 1.64<sup>***</sup> | (1.50-1.79) | 0.89<sup>*</sup> | (0.80-0.99) |
| **Measles' vaccine** |  |  |  |  |  |  |  |
| Not vaccinated | 86.7 | 13.3 | 0.898 | Ref. |  |  |  |
| Vaccinated | 86.7 | 13.3 |  | 1.01 | (0.92-1.10) |  |  |
| **Vitamin A supplement** |  |  |  |  |  |  |  |
| No | 87.2 | 12.8 | 0.055 | Ref. |  | Ref. |  |
| Yes | 86.2 | 13.8 |  | 1.09 | (1.00-1.20) | 1.1 | (1.00-1.22) |
| **Household Characteristics** |  |  |  |  |  |  |  |
| **Wealth index** |  |  |  |  |  |  |  |
| Poorest | 83.3 | 16.7 | <0.001 | Ref. |  | Ref. |  |
| Poorer | 85.4 | 14.6 |  | 0.85<sup>*</sup> | (0.75-0.97) | 0.9 | (0.79-1.03) |
| Middle | 87.5 | 12.5 |  | 0.71<sup>***</sup> | (0.62-0.82) | 0.77<sup>***</sup> | (0.67-0.89) |
| Richer | 88.3 | 11.7 |  | 0.66<sup>***</sup> | (0.57-0.77) | 0.79<sup>**</sup> | (0.66-0.93) |
| Richest | 90.2 | 9.8 |  | 0.54<sup>***</sup> | (0.47-0.64) | 0.69<sup>***</sup> | (0.55-0.86) |
| **Place of residence** |  |  |  |  |  |  |  |
| Urban | 91 | 9.0 | <0.001 | Ref. |  | Ref. |  |
| Rural | 85.5 | 14.5 |  | 1.71<sup>***</sup> | (1.49-1.97) | 1.13 | (0.98-1.30) |
| **Region** |  |  |  |  |  |  |  |
| Phnom Penh | 89.1 | 10.9 | 0.002 | Ref. |  | Ref. |  |
| Plain | 86.1 | 13.9 |  | 1.33<sup>*</sup> | (1.06-1.67) | 0.79<sup>*</sup> | (0.62-0.99) |
| Tonle Sap | 86 | 14.0 |  | 1.34<sup>**</sup> | (1.07-1.67) | 0.75<sup>*</sup> | (0.60-0.94) |
| Coastal | 89.8 | 10.1 |  | 0.93 | (0.72-1.19) | 0.53<sup>***</sup> | (0.41-0.69) |
| Mountain | 86.8 | 13.2 |  | 1.25 | (0.99-1.57) | 0.67<sup>**</sup> | (0.53-0.85) |
| **Drinking water** |  |  |  |  |  |  |  |
| Improved | 87.3 | 12.7 | 0.018 | Ref. |  | Ref. |  |
| Unimproved | 85.9 | 14.1 |  | 1.13<sup>*</sup> | (1.02-1.24) | 1.01 | (0.91-1.12) |
| **Toilet facility** |  |  |  |  |  |  |  |
| Improved | 90.4 | 9.6 | <0.001 | Ref. |  | Ref. |  |
| Non-Improved | 83.1 | 16.9 |  | 1.93<sup>***</sup> | (1.74-2.14) | 1.07 | (0.93-1.23) |
| **Survey years** |  |  |  |  |  |  |  |
| 2005 | 80.3 | 19.7 | <0.001 | Ref. |  | Ref. |  |
| 2010 | 85.0 | 15.0 |  | 0.72<sup>***</sup> | (0.63-0.82) | 0.74<sup>***</sup> | (0.64-0.85) |
| 2014 | 87.1 | 12.9 |  | 0.60<sup>***</sup> | (0.53-0.69) | 0.62<sup>***</sup> | (0.54-0.72) |
| 2022 | 93.8 | 6.2 |  | 0.27<sup>***</sup> | (0.23-0.31) | 0.32<sup>***</sup> | (0.26-0.38) |

To evaluate the potential effect modification of statistically significant associations by time and space, interaction models were estimated for each of the statistically significant factors from the adjusted analysis presented in **Table 3**. These factors were separately interacted with region and year. Almost all of these analyses resulted in interaction terms that were not statistically significant, indicating that the corresponding factors were associated with child diarrhea regardless of the region of Cambodia where a child lived or the year in which the data were collected. The one exception was the interaction between child age by residence and survey years, which resulted in a statistically significant interaction that was visualized with predicted probabilities (see **Fig 5**). The predicted probabilities indicate that children with increased age had a significantly lower probability of diarrhea in urban areas across Cambodia’s survey years than children living in rural areas.

**Fig 5.**
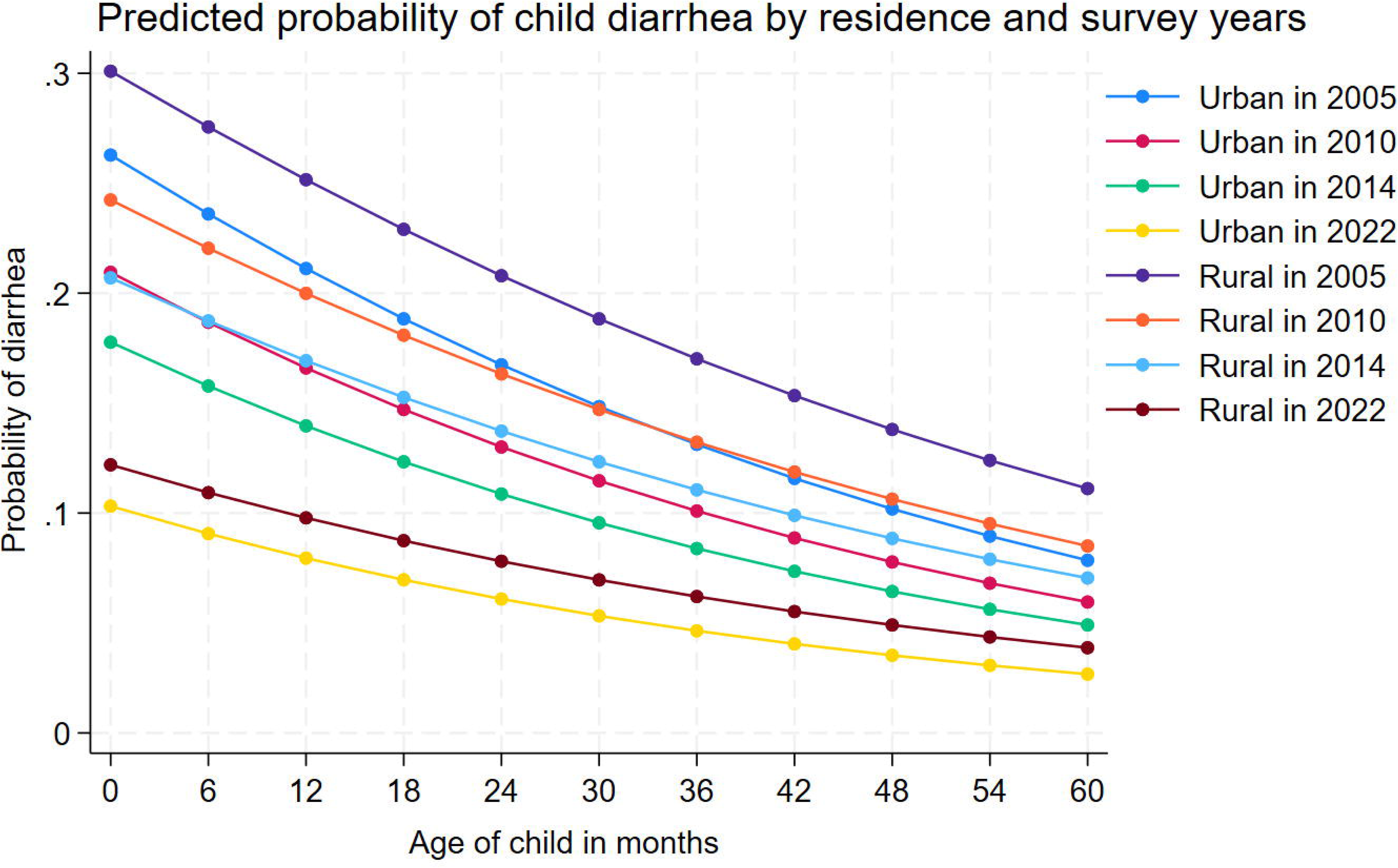
Predicted probability of child diarrhea in Cambodia by residences and survey years (N=29,742)

## Discussion

We have described the temporal and geospatial prevalence of diarrhea among children in Cambodia between 2005 and 2022, noting a significant decrease from 19.7% in 2005 to 15.0% in 2010 to 12.9%% in 2014 and 6.2% in 2022. This reduction in diarrhea burden corresponds with global efforts to reduce under-five mortality, and likely contributed to Cambodia’s achievement of Millennium Development Goal 4 (MDG-4) to reduce child mortality [6, 17, 18, 19]. This might be attributed to the efforts and initiative of the Royal Government of Cambodia (RGC), which has strengthened health facilities across the country, particularly in rural areas, improved the medical infrastructure, provided essential medical equipment and supplies, and improved safe drinking water and sanitation in rural area [6]. Recently, the RGC has prioritized child health and nutrition, closely monitored by the Prime Minister to ensure policy implementation and achieve Sustainable Development Goals 3 (SDGs-3) [20–22]. Cambodia has set ambitious goals within SDG-3, specifically aiming to reduce neonatal mortality to less than 12 per 1,000 live births and under-five mortality to less than 25 per 1,000 live births by 2030. These goals require continued focus on preventable childhood deaths, including diarrhea.

Our study found several factors associated with increased odds of having diarrhea among children aged 0–59 months in Cambodia. These include children born to mothers who smoke, mothers with limited access to healthcare services, and children aged 6-23 months. However, children were less likely to have diarrhea if their mothers were 25 or older, had at least a secondary education, were employed, or if the child was female, aged 36–60 months, lived in the richest wealth quintile, resided in a coastal region, or were surveyed in later years.

Children of mothers aged 25–34 years and 35–49 years have shown less occurrence of childhood diarrhea than young mothers. This is consistent with previous studies in Cambodia and India [9,16]. Likely, lower knowledge and experience of childcare and feeding among younger mothers can be a plausible causal explanation [9,17–19]. The odds of having diarrhea are lower among the children of working mothers, and similar findings have been identified in some recent studies in Cambodia, India, and Senegal [9,16,20]. Several studies document that the mother’s employment status may have an education, which will improve a child’s quality of living environment standards as well as improve hygienic practice, safe drinking water, and sanitation in the home during feeding and childcare [9,14,21,22]. Children of mothers completed secondary or higher education were found to be less likely than children whose mothers had no education to experience diarrhea. This is in line with findings from Cambodia [14,21]. This finding might be explained by the fact that highly educated mothers might have access to books and more education education programs that help to better knowledge and experience of childcare and feeding children [14,21].

Children from wealthier households were less likely to have diarrhea compared to those from poorer households, consistent with studies in Bangladesh, India, and Cambodia [15, 17, 22, 24]. This likely reflects increased access to nutrition and healthcare for children in wealthier households [15, 17, 22]. Higher income allows families to provide a more diverse diet, improved quality of care, and access to health services [25]. In this study, the age of the children significantly affected their risk of diarrhea. Children aged 36–47 months (AOR = 0.64; 95% CI: 0.52–0.79) and 48–60 months (AOR = 0.52; 95% CI: 0.42–0.66) had a lower odds ratio of having diarrhea compared to those aged 0–5 months. This finding aligns with similar studies [17, 22]. However, children aged 6–11 months (AOR = 2.05; 95% CI: 1.66–2.52) and 12–23 months (AOR = 1.68; 95% CI: 1.39–2.04) were more likely to have diarrhea compared to infants under five months. This aligns with research conducted in Northwest Ethiopia, India, and Cambodia [15, 17, 22, 23] and might be due to the developing immune system throughout childhood [22]. Females were less likely to have diarrhea than males, which may reflect a natural predisposition of males to develop diarrhea more frequently than females [8].

Children of smoking mothers were 1.61 times more likely to have diarrhea compared to children of non-smoking mothers. A narrative review focused on smoking increases the risk of infectious diseases, which found that investigators found that smoking was the leading risk factor in a reduced immune response [23]. Children of mothers having a problem accessing healthcare services were 1.20 times more likely to have diarrhea compared to children of mothers not having a problem. This aligns with findings from 31 countries in sub-Saharan Africa [24]. For women, access to medical care is one buffer against poor health. However, disadvantaged women experience limited access to care and are more likely to have an unplanned pregnancy, partly reflecting inadequate preconception care and family planning. They are also more likely to start prenatal care later than advantaged peers, deliver in hospitals with lower quality-of-care indicators, and are less likely to know about childcare [24]. Lastly, children living in rural areas had AOR=1.13 times more likely to have diarrhea than in urban areas. This result was in line with the study conducted in India and Cambodia [9,16]. This could be due to limited access to healthcare and sanitation facilities in rural areas [14,21,22]. Thus, compared to children living in households with unimproved drinking water, children from households with unimproved toilets were found not significantly associated with diarrhea. Inconsistent with other studies, Improved sanitation facilities were found in a supporting multicounty WASH-intervention study to reduce the risk of children from infection disease [25,26].

## Limitation

This study used pooled data from Cambodia Demographic and Health surveys from 2005, 2010, 2014, and 2022. This enabled us to describe the temporal and geographical trends in diarrhea among children aged 0–59 months using nationally representative data [14,15]. Likewise, we were able to examine factors associated with diarrhea among children throughout Cambodia. We acknowledge that the results from this study may reflect the present prevalence of diarrhea among children in the country by comparing the associated factors with diarrhea among children to another DHS survey. It is hoped that the evidence will help the Cambodian Ministry of Health’s Child Health program, as well as health promotion and social determinants, to plan interventions to reduce child morbidity and mortality due to diarrhea. However, CDHS collected data as a cross-sectional study, so examining causality in these relationships was impossible. Also, the outcome variable was collected from self-reporting from mothers or caregivers to define the presence of diarrhea among eligible children in the past two weeks before data collection, making our analysis prone to recall bias. However, this recall bias appears non-differential since the DHS survey uses the standard questionnaire recommended by UNICEF/WHO [27]. As a secondary data analysis, several known etiologies of diarrhea, such as parasites, viruses, bacteria, and common factors for diarrhea. These included children’s malnutrition, history of illness, antibiotic prescription, type of roofing material, household types of cooking, season effect, and protective vaccination against diarrhea, which were not included in this study due to data limitations.

## Conclusion

Our findings suggest that the relatively high prevalence of diarrhea among children aged 0–59 months in Cambodia declined slightly over the 20 years between 2005 and 2022. Diarrhea among children continues to be a significant public health concern in Cambodia. This study indicated that the factors associated with increased odds of having diarrhea among children aged 0–59 months in Cambodia, including children of smoking mothers, mothers having a problem accessing healthcare services, children aged 6-23 months, and children living in rural areas. However, children born to mothers aged 25 or above, mothers who completed at least secondary education, maternal employment, females, children aged 36–60 months, children living in the richest wealth quintiles, coastal regions, and survey years were less likely to have diarrhea. Public health practitioners and policymakers should consider demographic groups represented by these characteristics to produce better-targeted interventions to address diarrhea among children in Cambodia.

## Data Availability

The CDHS data are publicly available from the website at (URL: https://www.dhsprogram.com/data/available-datasets.cfm).

## Acknowledgments

The authors would like to thank DHS-ICF, who approved the data used for this paper.

## Data Availability Statement

Our study used 2005, 2010, 2014, and 2021-22 Cambodia Demographic and Health Survey (CDHS) datasets. The DHS data are publicly available from the website at (URL: https://www.dhsprogram.com/data/available-datasets.cfm).

## Abbreviations

ACS: American Cancer Society
AOR: adjusted odds ratio
CDHS: Cambodia Demographic Health Survey
EA: enumeration areas
PPS: probability proportional to size

## Supporting information

**Table S1.** Results of checking multicollinearity using Variance Inflation Factor.

| **Variable** | **VIF** |
| --- | --- |
| Wealth quintile | 2.16 |
| Type of toilet facility | 2.11 |
| Mother age | 1.50 |
| Survey years | 1.47 |
| Mother education | 1.45 |
| Place of residence | 1.32 |
| Geographical region | 1.24 |
| Smoking | 1.10 |
| Maternal empyloment | 1.02 |
| Sex of child | 1.00 |
| Mean VIF | 1.42 |

## References

1. WHO. World Health Organization. Diarrhoeal disease; Key facts 2024 [7 March 2024]. Available from: https://www.who.int/news-room/fact-sheets/detail/diarrhoeal-disease.

2. WHO. Global Health Estimates 2020: Disease burden by Cause, Age, Sex, by Country and by Region, 2000–2019. Geneva, Switzerland: World Health Organization, 2020.

3. National Institute of Public Health, National Institute of Statistics, and ORC Macro. Cambodia Demographic and Health Survey 2005. Phnom Penh, Cambodia and Calverton, Maryland, USA: National Institute of Public Health, National Institute of Statistics and ORC Macro. 2005. Available online: https://dhsprogram.com/pubs/pdf/FR185/FR185%5BApril-27-2011%5D.pdf. 2006.

4. National Institute of Statistics, Directorate General for Health, and ICF International. Cambodia Demographic and Health Survey 2014. Phnom Penh, Cambodia, and Rockville, Maryland, USA: National Institute of Statistics, Directorate General for Health, and ICF International. Available online: https://dhsprogram.com/pubs/pdf/fr312/fr312.pdf. 2015.

5. National Institute of Statistics, Directorate General for Health, and ICF Macro. Cambodia Demographic and Health Survey 2010. Phnom Penh, Cambodia and Calverton, Maryland, USA: National Institute of Statistics, Directorate General for Health, and ICF Macro. 2010. Available online: https://dhsprogram.com/pubs/pdf/Fr249/Fr249.pdf. 2011.

6. National Institute of Statistics PP, Cambodia, Ministry of Health Phnom Penh, Cambodia, The DHS Program, ICF, Rockville, Maryland, USA. Cambodia Demographic and Health Survey 2021–22. Phnom Penh, Cambodia, and Rockville, Maryland, USA: NIS, MoH, and ICF. 2022. Available from: https://dhsprogram.com/pubs/pdf/FR377/FR377.pdf.

7. Sharrow D, Hug L, You D, Alkema L, Black R, Cousens S, et al. Global, regional, and national trends in under-5 mortality between 1990 and 2019 with scenario-based projections until 2030: a systematic analysis by the UN Inter-agency Group for Child Mortality Estimation. Lancet Glob Health. 2022;10(2):e195–e206. doi: 10.1016/s2214-109x(21)00515-5. PubMed PMID: 35063111; PubMed Central PMCID: PMCPMC8789561.

8. Arifin H, Rakhmawati W, Kurniawati Y, Pradipta RO, Efendi F, Gusmaniarti G, et al. Prevalence and determinants of diarrhea among under-five children in five Southeast Asian countries: Evidence from the demographic health survey. Journal of Pediatric Nursing. 2022.

9. Pisey V, Banchonhattakit P, Laohasiriwong W. The association of socio-demographic and environmental factors on childhood diarrhea in Cambodia. F1000Research. 2020;9.

10. Sarker AR, Sultana M, Mahumud RA, Sheikh N, Van Der Meer R, Morton A. Prevalence and health care–seeking behavior for childhood diarrheal disease in Bangladesh. Global pediatric health. 2016;3:2333794X16680901.

11. STATA. StataCorp. 2023. Stata Statistical Software: Release 18. College Station, TX: StataCorp LLC. 2023.

12. Desktop A. Release 10.3.(2020). ESRI (Environmental Systems Research Institute), Redlands, CA.

13. Cunningham SA, J. Muir, J. A. “Data Cleaning”. The Cambridge Handbook of Research Methods and Statistics for the Social and Behavioral Sciences: Volume 1: Building a Program of Research. Cambridge Handbooks in Psychology. Cambridge: Cambridge University Press; 2023. p. 443–67. 2023. doi: 10.1017/9781009010054.022.

14. Um S, Cope MR, Muir JA. Child anemia in Cambodia: A descriptive analysis of temporal and geospatial trends and logistic regression-based examination of factors associated with anemia in children. PLOS Glob Public Health. 2023;3(9):e0002082. Epub 20230915. doi: 10.1371/journal.pgph.0002082. PubMed PMID: 37713392; PubMed Central PMCID: PMCPMC10503718.

15. Um S, Sopheab H, Yom A, Muir JA. Anemia among pregnant women in Cambodia: A descriptive analysis of temporal and geospatial trends and logistic regression-based examination of factors associated with anemia in pregnant women. PLoS One. 2023;18(12):e0274925. Epub 20231207. doi: 10.1371/journal.pone.0274925. PubMed PMID: 38060474; PubMed Central PMCID: PMCPMC10703242.

16. Ghosh K, Chakraborty AS, Mog M. Prevalence of diarrhoea among under five children in India and its contextual determinants: A geo-spatial analysis. Clinical Epidemiology and Global Health. 2021;12:100813.

17. Nilima N, Kamath A, Shetty K, Unnikrishnan B, Kaushik S, Rai SN. Prevalence, patterns, and predictors of diarrhea: a spatial-temporal comprehensive evaluation in India. BMC public health. 2018;18:1–10.

18. Edwin P, Azage M. Geographical variations and factors associated with childhood diarrhea in Tanzania: a national population based survey 2015-16. Ethiopian journal of health sciences. 2019;29(4).

19. Ghasemi AA, Talebian A, Masoudi Alavi N, Moosavi G. Knowledge of mothers in management of diarrhea in under-five children, in kashan, iran. Nurs midwifery stud. 2013;1(3):158–62.

20. Thiam S, Diène AN, Fuhrimann S, Winkler MS, Sy I, Ndione JA, et al. Prevalence of diarrhoea and risk factors among children under five years old in Mbour, Senegal: a cross-sectional study. Infect Dis Poverty. 2017;6(1):109. Epub 20170706. doi: 10.1186/s40249-017-0323-1. PubMed PMID: 28679422; PubMed Central PMCID: PMCPMC5499039.

21. Um S, Vang D, Pin P, Chau D. Trends and determinants of acute respiratory infection symptoms among under-five children in Cambodia: Analysis of 2000 to 2014 Cambodia demographic and health surveys. PLOS Glob Public Health. 2023;3(5):e0001440. Epub 20230503. doi: 10.1371/journal.pgph.0001440. PubMed PMID: 37134089; PubMed Central PMCID: PMCPMC10155954.

22. Vong P, Banchonhattakit P, Sim S, Pall C, Dewey RS. Unhygienic stool-disposal practices among mothers of children under five in Cambodia: Evidence from a demographic and health survey. PloS one. 2021;16(7):e0249006. Epub 20210701. doi: 10.1371/journal.pone.0249006. PubMed PMID: 34197455; PubMed Central PMCID: PMCPMC8248716.

23. Jiang C, Chen Q, Xie M. Smoking increases the risk of infectious diseases: A narrative review. Tob Induc Dis. 2020;18:60. Epub 20200714. doi: 10.18332/tid/123845. PubMed PMID: 32765200; PubMed Central PMCID: PMCPMC7398598.

24. Adedokun ST, Yaya S. Factors influencing mothers’ health care seeking behaviour for their children: evidence from 31 countries in sub-Saharan Africa. BMC Health Serv Res. 2020;20(1):842. Epub 20200907. doi: 10.1186/s12913-020-05683-8. PubMed PMID: 32894107; PubMed Central PMCID: PMCPMC7487813.

25. Naz L, Ghimire U. Unimproved water, sanitation, and hygiene (WASH) and common childhood illness in Myanmar: evidence from a nationally representative survey. 2020.

26. Gera T, Shah D, Sachdev HS. Impact of Water, Sanitation and Hygiene Interventions on Growth, Non-diarrheal Morbidity and Mortality in Children Residing in Low- and Middle-income Countries: A Systematic Review. Indian Pediatr. 2018;55(5):381–93. Epub 20180209. PubMed PMID: 29428924.

27. Rego R, Watson S, Alam MAU, Abdullah SA, Yunus M, Alam IT, et al. A comparison of traditional diarrhoea measurement methods with microbiological and biochemical indicators: A cross-sectional observational study in the Cox’s Bazar displaced persons camp. EClinicalMedicine. 2021;42:101205. Epub 20211120. doi: 10.1016/j.eclinm.2021.101205. PubMed PMID: 34849477; PubMed Central PMCID: PMCPMC8608865.

